# Humanization of Care: A Geospatial Analysis of Key Indicators of Quality and Safety of Health Care and Service in the Centre Region, Cameroon

**DOI:** 10.1101/2024.11.11.24317125

**Authors:** Fabrice Zobel Lekeumo Cheuyem, Adidja Amani, Brian Ngongheh Ajong, Lionel Berthold Keubou Boukeng, Christian Mouangue, Martine Golda Mekouzou Tsafack, Esther Andriane Bitye Bi Mvondo, Ariane Nouko, Claude Axel Minkandi, Christelle Sandrine Ngos, Edwige Omona Guissana, Raissa Katy Noa Otsali, Djoulay-Hatou, Adam Yaya, Pamela Sonfack, Florence Kissougle Nkongo

## Abstract

**Background:** Integrating the principles of humanized care into healthcare systems is critical to promoting optimal well-being throughout the patient care process. In a context of limited resources, improving the quality of care and health services means identifying priority sites to optimize cost-effectiveness. The objective was to measure key indicators of quality and safety of care in health facilities (HFs) and to map priority health districts (HDs) for effective implementation of high-impact interventions in the Centre Region.

**Methods:** We conducted a descriptive cross-sectional analysis using retrospective data from 2018 to 2022 from 32 HDs comprising the Centre Region were extracted in April 2023. Data were retrieved from the national database using DHIS 2 and checked for completeness. The health indicators assessed were those defined in the Cameroon Health Sector Strategy for 2020-2030.

**Results:** The density of the qualified HCWs was 13.7 per 10 000 inhabitants in 2022. The density of doctors in rural areas has fallen sharply over the five-year study period, with a decline of almost 20% by 2020 in favor of urban areas. The Odza HD had the highest inpatient mortality rate over the five-year study period (49%). In rural areas, the Esse HD had the highest mortality rate (32.4%). Esse (18.8%) and Odza (6.7%) HDs had the highest perioperative mortality rates in rural and urban areas, respectively. Most of the urban HDs (5/8=62.5%) had a neonatal mortality rate higher than the regional median (RM=3.1). Half of the rural HDs had a maternal mortality ratio higher than the regional median (RM=118 per 1000 live births). The HDs of Mbandjock (357/1,000) and Mvog-Ada (285/1,000) were the most affected in rural and urban areas respectively. More than two thirds of urban HDs recorded a proportion of free malaria treatment cases below the regional median (14%).

**Conclusions:** Efforts to humanize healthcare in the Centre Region are hindered by persistent disparities in care quality and inconsistent policy implementation. Addressing these challenges requires timely implementation of a National Strategic Plan to Improve the Quality of Healthcare and Service Delivery, emphasizing on capacity building through training and supervision.

## Background

In the healthcare systems, humanization of care is an essential component in promoting well-being during the care process as hospital environments often create emotional imbalances due to the suffering and anxiety experienced by patients and their families [1,2]. Humanization of health services seeks to enhance care quality ensuring comprehensive patient support without causing financial strain throughout their lives [3,4].

Humanized care is an enlightening guide and support to practice for its benefit in cultivating an awareness of care and establishing a strong and lasting therapeutic relationship. Nursing, therefore, is rooted in humanistic values aimed at meeting patients’ needs and improve their quality of life [5]. This humanized care process requires qualified, well-trained and available healthcare workers (HCWs) to effectively address people’s health problems.

The third objective of Sustainable Development Goals (SDGs) by 2030 aims to reduce the global maternal mortality ratio to less than 70 per 100,000 live births and eliminate all preventable infant and under-five mortality, with all countries aiming to reduce neonatal mortality to no more than 12 per 1,000 live births and under-five mortality to no more than 25 per 1,000 live births [6,7].

Although the proportion of doctors in Cameroon is twice (1.9 per 1,000 people) the minimum recommended by the World Health Organization, the country’s health statistics remain concerning. Cameroon continues to experience limited reductions in under-five mortality compared to other nations [8]. Providing safe, high-quality care is one of the components of humanized care, as it facilitates patient recovery and quality of life and has a positive impact on the intangible costs of care [9].

In the Health Sector Strategy for 2020-2030, health indicators related to the quality of care have been defined. Such indicators include the maternal mortality rate, the neonatal mortality rate, the infant mortality rate, the child mortality rate, and the direct maternal in-hospital mortality rate. Targets have been assigned to these key indicators, and they will be evaluated once the strategies for improving health in Cameroon have been implemented [10].

To improve the quality of care and reduce under-five mortality, the national policy in the context of universal health coverage provides free treatment of simple and severe forms of malaria in children under 5 years of age in health facilities (HFs), using molecules subsidized by the government and its donors (artemether & lumefantrine and artesunate injection) [11]. The objective of our study was to assess key indicators of quality and safety of care in HFs and to map priority health districts (HDs) for effective implementation of high-impact interventions in the Centre Region.

## Methods

### Study Type

The present study was a descriptive cross-sectional analysis.

### Study Period

The online database was consulted in April 2023 to retrieve health data from 2018 to 2022 (five-year period).

### Study Site

This evaluation covered the 32 HDs of the Centre Region. They were grouped into the urban area (Yaounde), which includes the HDs of Biyem-Assi, Cite-Verte, Efoulan, Mvog-Ada, Djoungolo, Nkolndongo, Nkolbisson and Odza, and the rural area, represented by the remaining 24 HDs (Figure 1).

**Fig. 1.**
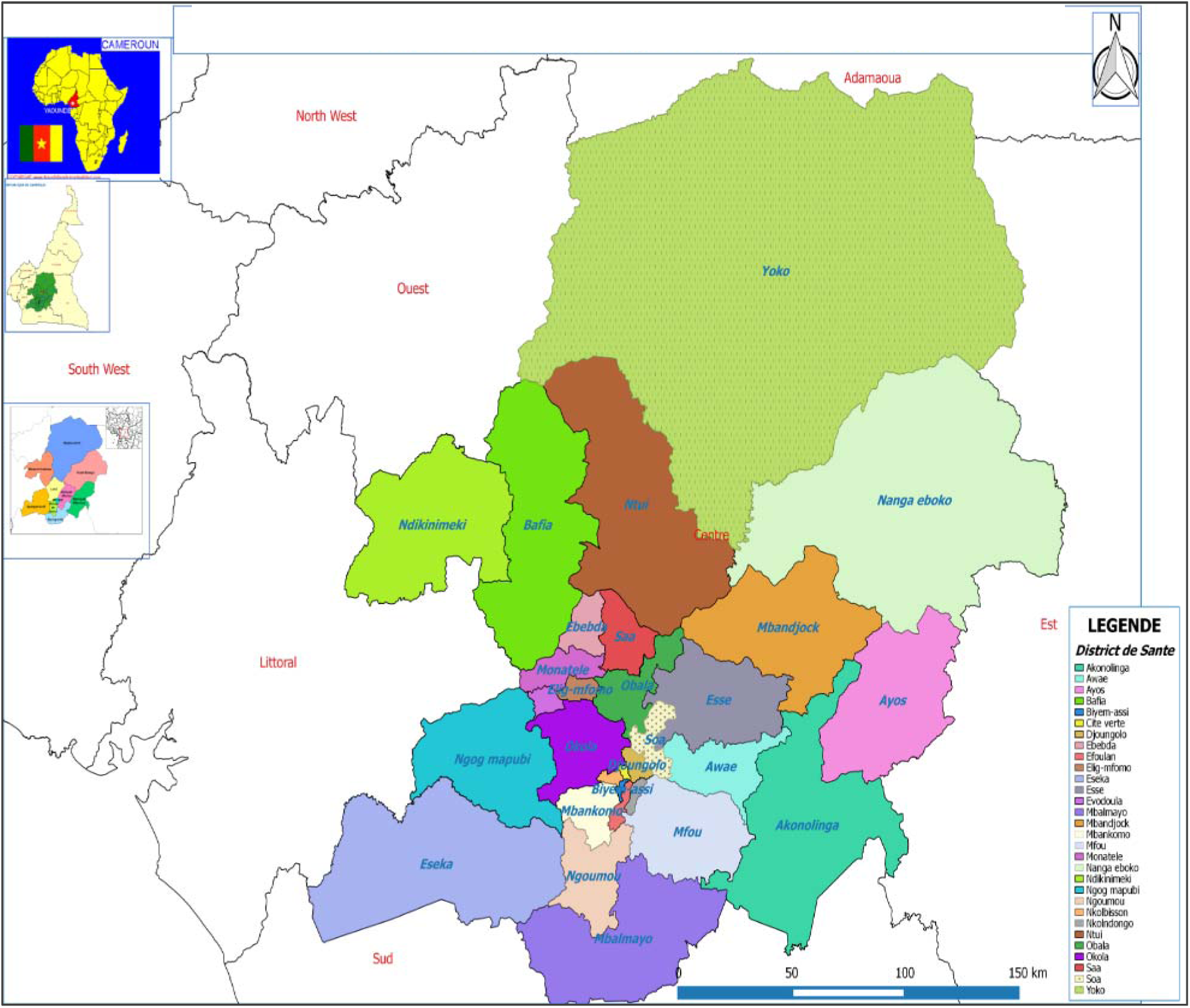
Map of Yaounde Health Districts [12]

### Data Sources and Processing

Data were retrieved from the national database using the DHIS 2 pivot table tool in a Microsoft Office Excel 2016 file. Following national guidelines, this tool is updated monthly with data from the HFs in each HD. After extraction, the data were checked for accuracy and completeness before data entry. Any data discrepancies identified were resolved by contacting the regional data manager or district medical officer directly. Data collected included peri-operative, neonatal and maternal mortality rates, and uptake of free malaria treatment for children under five in HFs in the Centre Region. Data collected are those defined in the Health Sector Strategy for 2020-2030 [10].

### Operational Definition

Qualified HCWs include Physicians, other clinicians, state-registered nurses, and midwives [13]. The perioperative mortality rate is the rate of death (from all causes) before hospital discharge among patients who have undergone one or more surgical procedures during their hospital stay. The neonatal mortality rate is the probability that a child born in a given place in a given year or period will die in the first 28 days of life, expressed per 1000 live births. The maternal mortality ratio refers to the annual number of deaths among women, due to causes related to, or exacerbated by pregnancy or its management (excluding accidental or fortuitous causes), occurring during pregnancy or childbirth or in the 42 days following the end of pregnancy, whatever the duration or type of pregnancy, expressed per 1000 live births, over a specified period [14].

### Data Analysis

Data were analyzed using R Statistics software version 4.2.3 and maps were produced using QGIS Desktop software version 3.30.1. The HDs were classified into four priority categories, with red representing the highest priority and dark green the lowest. The defined categories corresponded to the four quartiles of each assessed health indicator. Logistic regression was used to identify the most vulnerable areas that will require more attention. The dependent variable was the indicator being assessed and the independent variable was the environment to which the district belonged. Regional thresholds represent the median for each health indicator measured. The significance level was set at 5% and the confidence level at 95%.

## Results

### Human Resource for Health Analysis

The analysis of health care and service availability index for the Centre Region revealed a density of the qualified HCWs of 13.7 per 10,000 inhabitants in 2022 (Table 1).

**Table 1.** Indices for the availability of health care and services in the Centre Region in 2022.

| Indicator | Target | Value |
| --- | --- | --- |
| **Infrastructure** |  |  |
| Density of health facilities<sup>1</sup> | 2 | 1.4 |
| Hospital bed<sup>1</sup> | 25 | 20.1 |
| **Healthcare worker** |  |  |
| Qualified staff<sup>1</sup> | 23 | 13.7 |
| **Use of services** |  |  |
| Use (outpatient consultations per person/year) | 5 | 0.5 |

<sup>1</sup> Value per 10 000 inhabitants

This variable shows a downward trend in urban areas from 2019 to 2022. However, since 2018 in rural areas, it has risen sharply and stabilized at around 15 per 10,000 inhabitants. As for the density of nursing staff, it has increased overall over the last five years in both zones. However, a break in this trend was observed in 2022 (Figure 2,3).

**Fig. 2.**
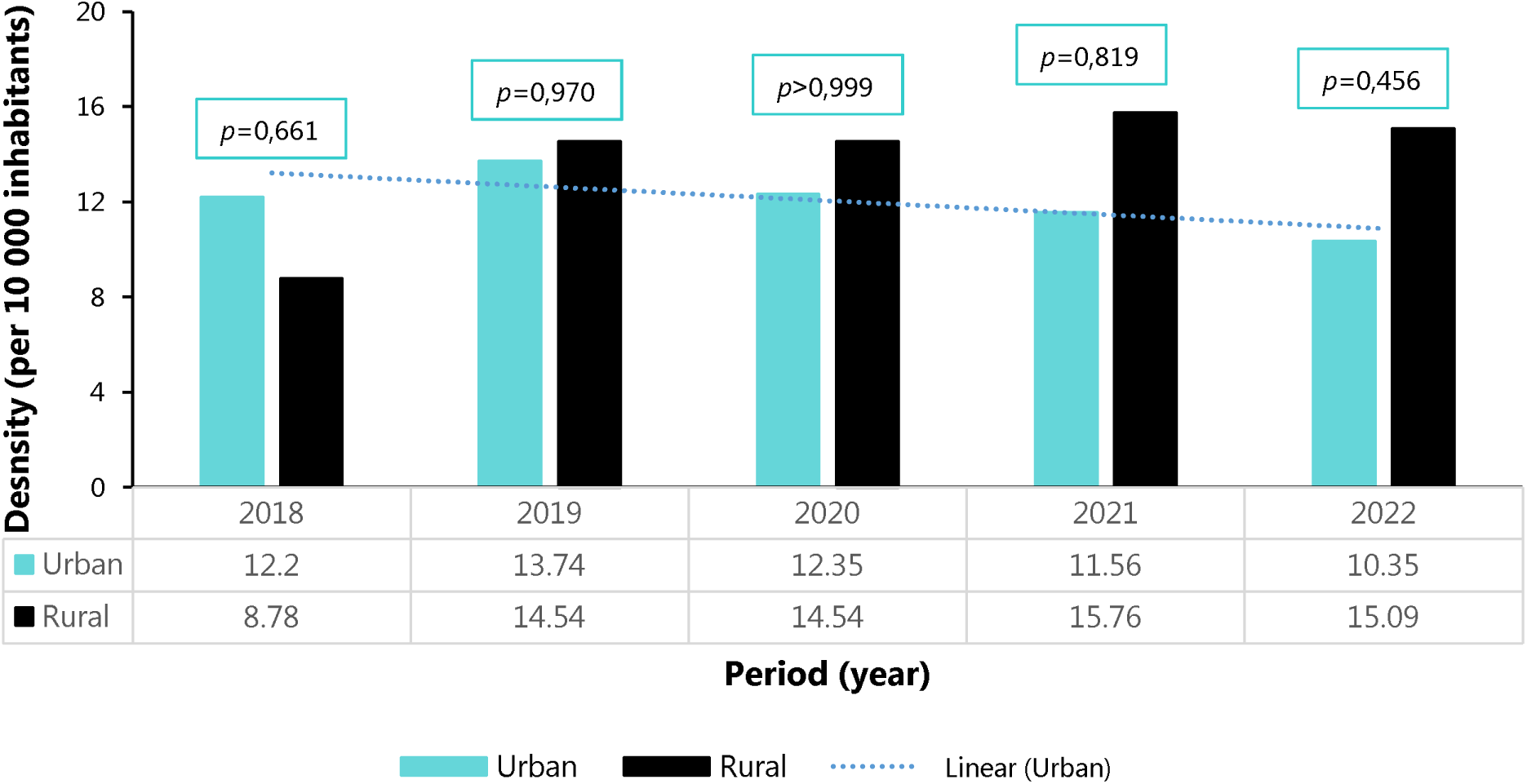
Density of qualified healthcare workers in urban and rural health facilities in the Centre Region from 2018 to 2022

**Fig. 3.**
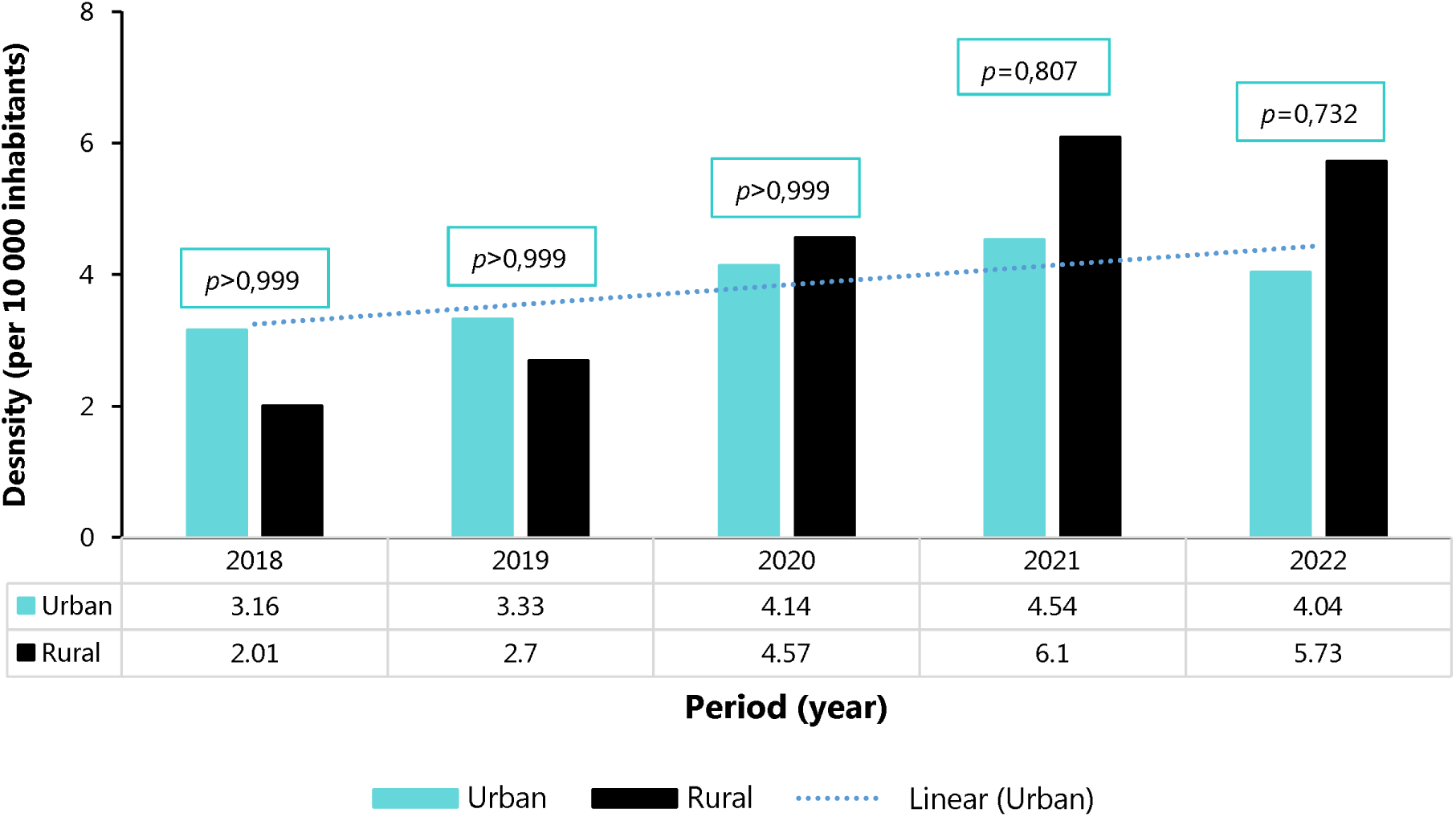
Density of nurses in urban and rural health facilities in the Centre Region from 2018 to 2022

The density of doctors in rural areas has fallen sharply over the five-years study period, with a decline of almost 20% by 2020 in favor of urban areas (Figure 4).

**Fig. 4.**
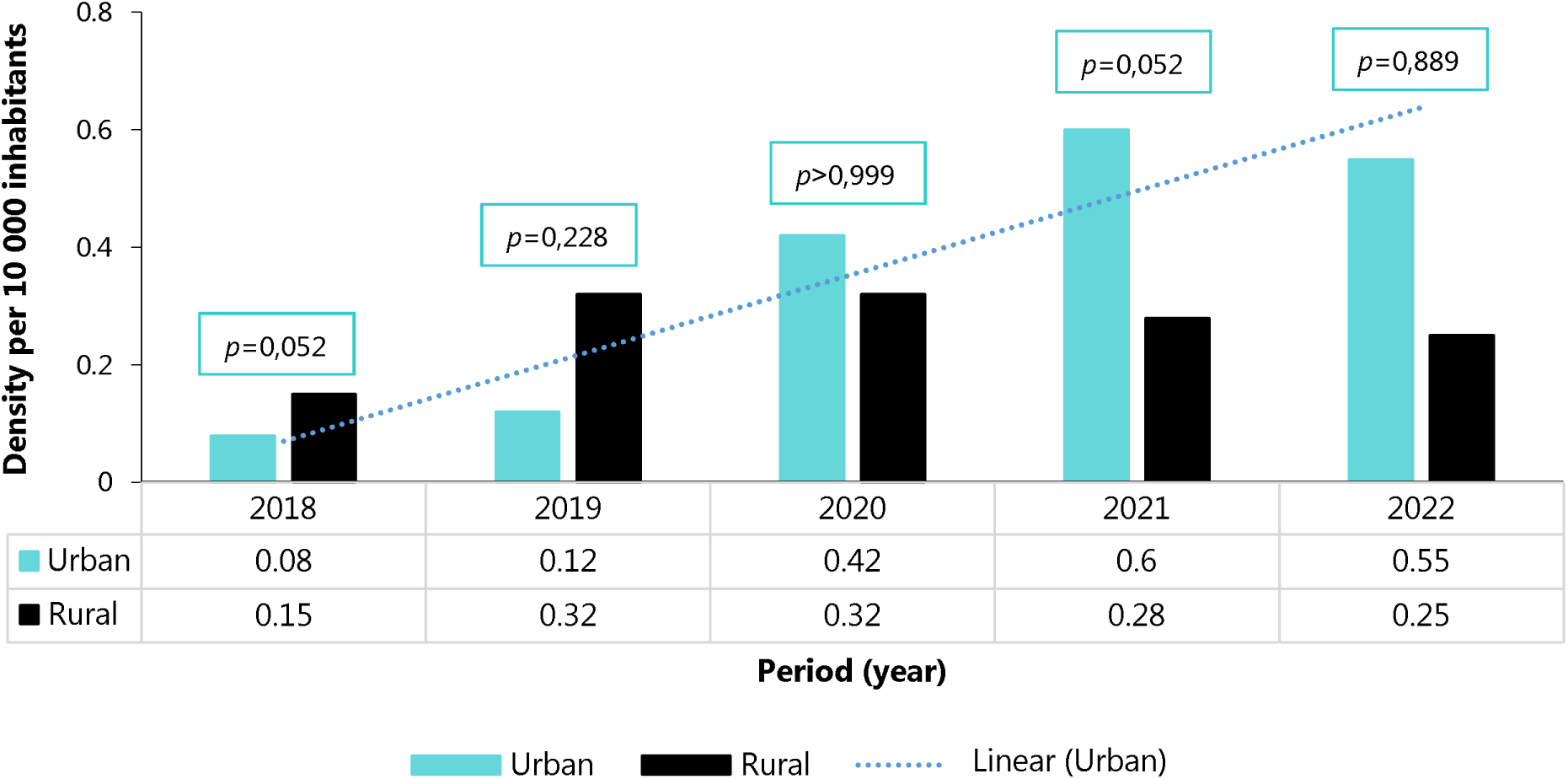
Density of doctors in urban and rural health facilities in the Centre Region from 2018 to 2022

The density of qualified HCWs midwives increase in both urban and rural areas over five-years study period. However, the trend was slightly higher in rural HDs than in urban areas (Figure 5).

**Fig. 5.**
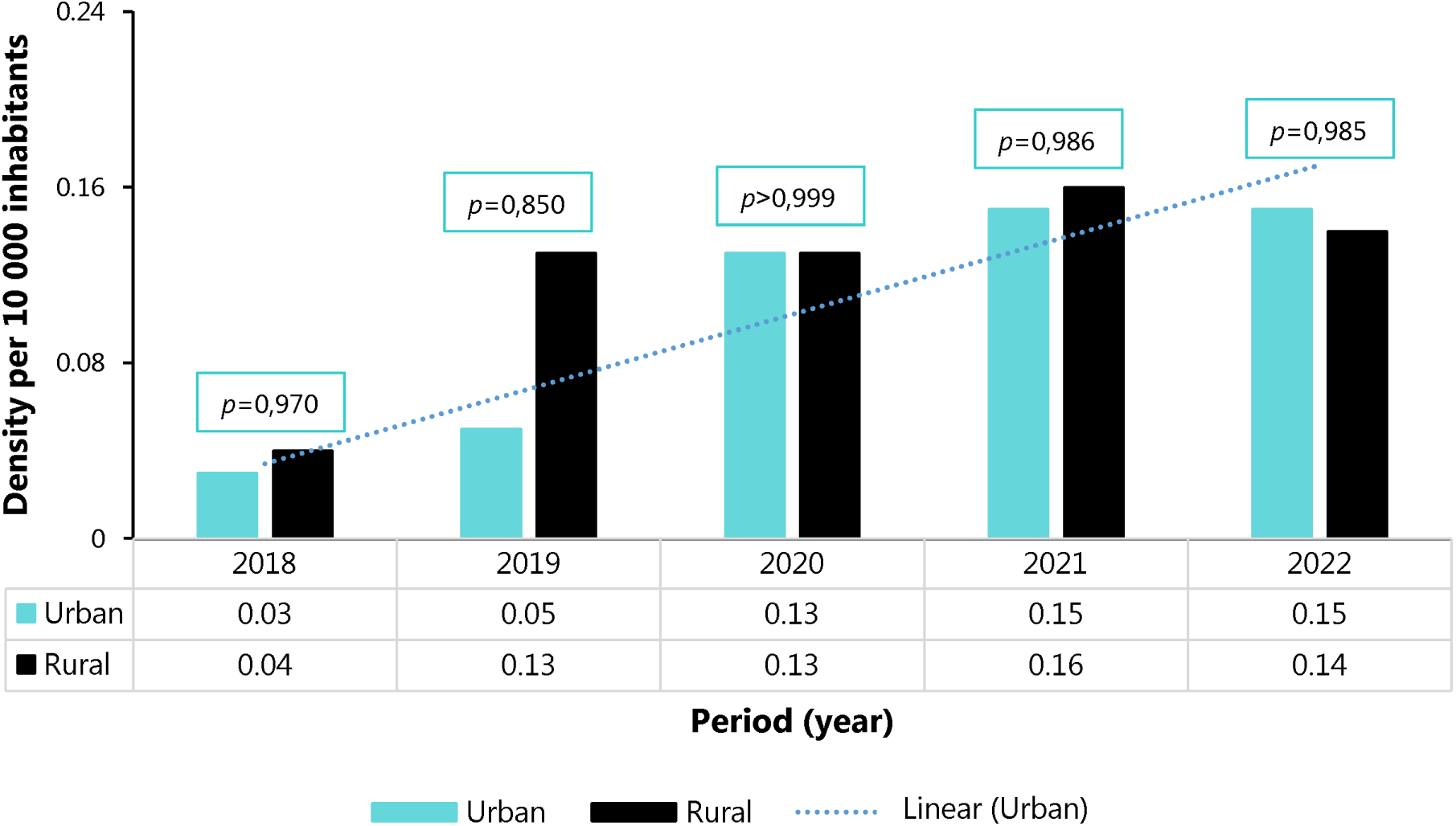
Density of midwives in urban and rural health facilities in the Centre Region from 2018 to 2022

### Inpatient Mortality

The Odza HD had the highest inpatient mortality rate in the Centre Region over the five-years study period (49%). In addition to this HD, three others had mortality rates above the regional median (15.4%): Cite-Verte, Mvog-Ada and Biyem-Assi. In rural areas, the Esse HD had the highest mortality rate (32.4%) (Figure 6).

**Fig. 6.**
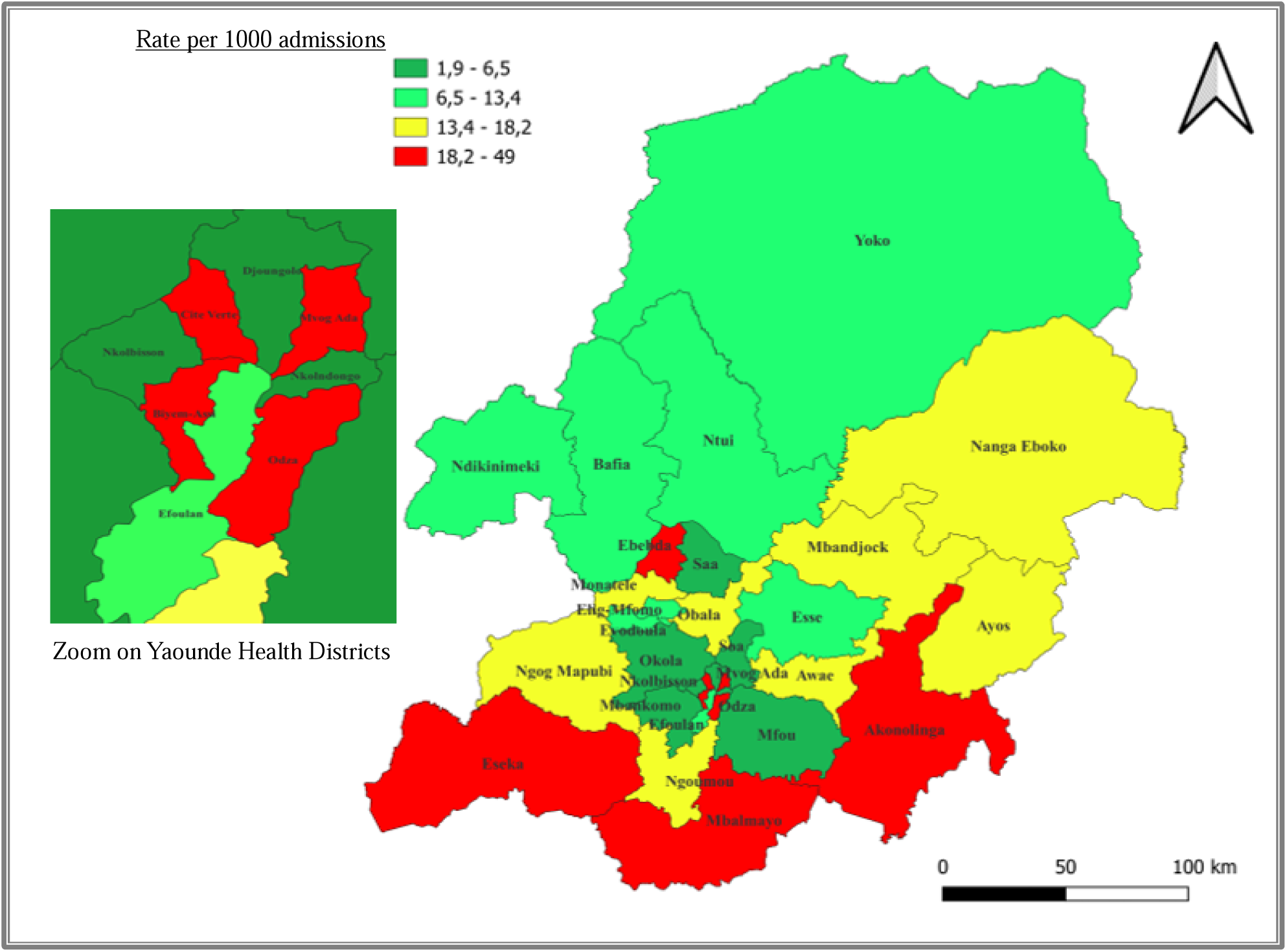
Mapping of inpatient mortality in the Health Districts of the Centre Region from 2018 to 2022

Regarding inpatient mortality, the univariate analysis did not show any significant difference between urban and rural areas compared to the regional median mortality rate (RM=13.5 per 1000 admissions) (Table 2).

**Table 2.**
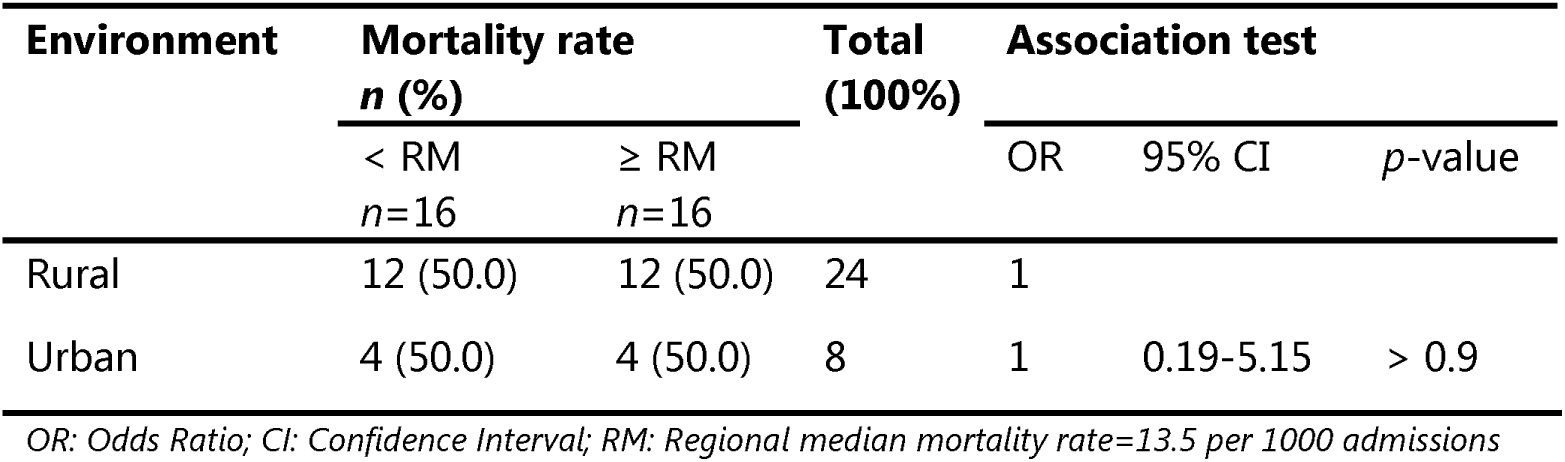
Bivariate analysis of the excess risk of inpatient mortality by type of environment in the Centre Region (*n=*32)

### Perioperative Mortality

Analysis of perioperative mortality in the Centre Region showed that over the five-years study period, Esse (18.8%) and Odza (6.7%) HDs had the highest perioperative mortality rates in rural and urban areas, respectively, (Figure 7).

**Fig. 7.**
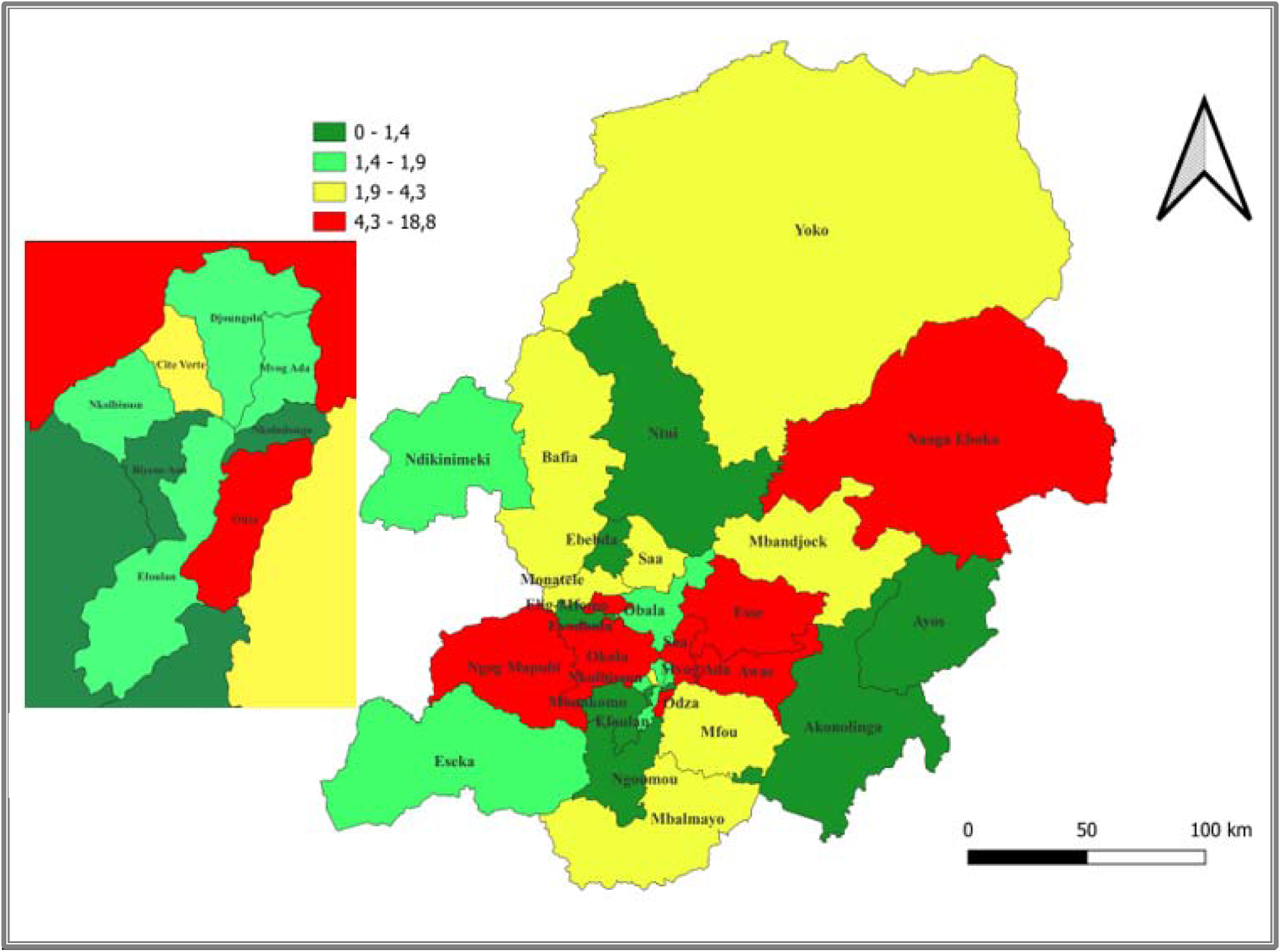
Mapping of perioperative mortality in the Health Districts of the Centre Region from 2018 to 2022

More than half of the rural HDs had a mortality rate above the regional median, with the risk of perioperative mortality 4.2 times higher than in urban areas (*p*=0.117) (Table 3).

**Table 3.** Bivariate analysis of the excess risk of perioperative mortality according to type of environment in the Centre Region’s Health Districts (*n*=32)

| Environment | Mortality rate<br>*n* (%) |  | Total<br>(100%) | Association test |  |  |
| --- | --- | --- | --- | --- | --- | --- |
|  | < RM<br>*n*=16 | ≥ RM<br>*n*=16 |  | OR | 95% CI | *p*-value |
| Urban | 6 (75.0) | 2 (25.0) | 8 | 1 |  |  |
| Rural | 10 (41.7) | 14 (58.3) | 24 | 4.20 | 0.78 - 33.0 | 0.117 |

OR: Odds Ratio; CI: Confidence Interval; RM: Regional median mortality rate=1,95%

### Neonatal Mortality

Most of the urban HDs in the Centre Region (5/8=62.5%) had a neonatal mortality rate higher than the regional median (RM=3.1%), with a mortality risk 1.41 times higher than in rural health districts (*p*=0.68) (Table 4).

**Table 4.** Bivariate analysis of excess risk of neonatal mortality in the DS of the Centre according to type of environment (*n*=32)

| Environment | Mortality rate<br>*n* (%) |  | Total<br>(100%) | Association test |  |  |
| --- | --- | --- | --- | --- | --- | --- |
|  | < RM<br>*n*=14 | ≥ RM<br>*n*=18 |  | OR | 95% CI | *p*-value |
| Rural | 11 (45.8) | 13 (54.2) | 24 | 1 |  |  |
| Urban | 3 (37.5) | 5 (62.5) | 8 | 1.41 | 0.28-8.16 | 0.681 |

OR: Odds Ratio; CI: Confidence Interval; RM: Regional median mortality rate=3.1 per 1000 live births

The highest rates were recorded in the health districts of Cite-Verte (9.4%) and Ndikiniméki (8%) (Figure 8).

**Fig. 8.**
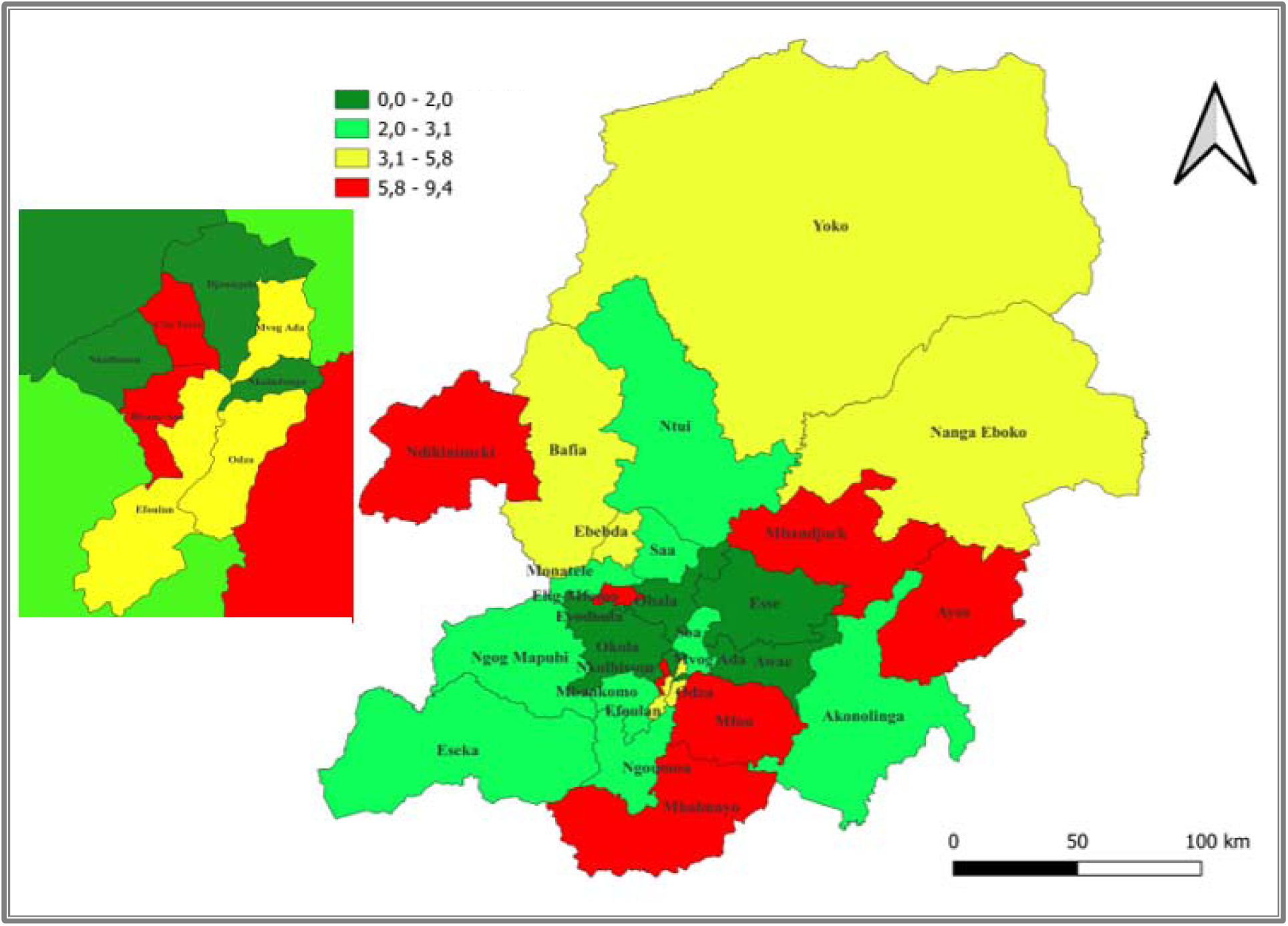
Neonatal mortality in the Health Districts of the Centre Region from 2018 to 2010

### Maternal Mortality

Half of the rural HDs had a maternal mortality ratio higher than the regional median (118 per 1000 live births). The HDs of Mbandjock (357/1,000) and Mvog-Ada (285/1,000) were the most affected in rural and urban areas respectively (Figure 9, Table 5).

**Fig. 9.**
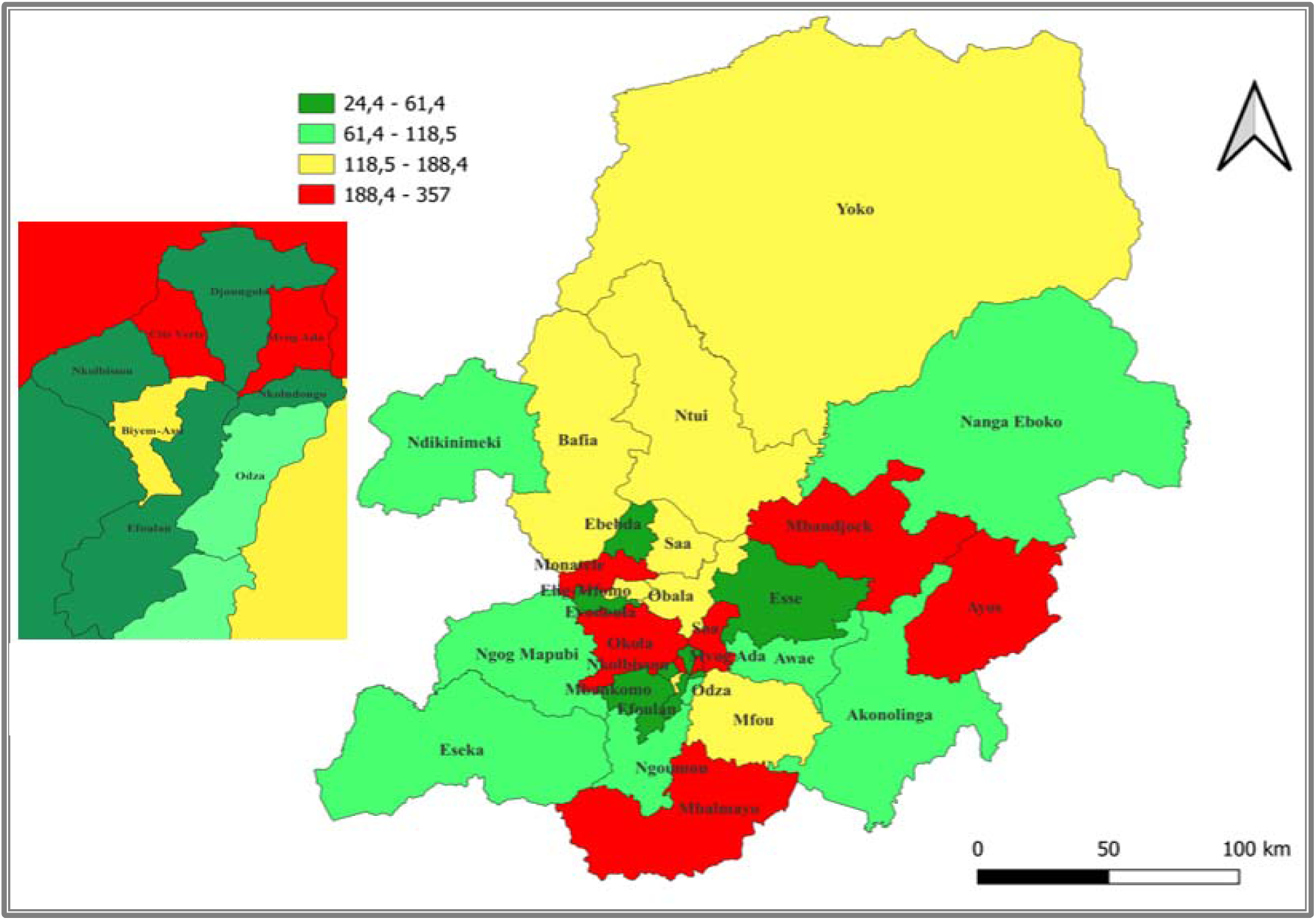
Mapping of in-hospital maternal mortality in the Health Districts of the Centre Region from 2018 to 2022

**Table 5.** Bivariate analysis of excess risk of in-hospital maternal mortality ratio by type of environment in the Centre Region (*n*=32)

| Environment | Mortality rate<br>*n* (%) |  | Total<br>(100%) | Association test |  |  |
| --- | --- | --- | --- | --- | --- | --- |
|  | < RM<br>*n*=16 | ≥ RM<br>*n*=16 |  | OR | 95% CI | *p*-value |
| Urban | 5 (62.5) | 3 (37.5) | 8 | 1 |  |  |
| Rural | 11 (45.8) | 13 (54.2) | 24 | 1.41 | 0.28-8.16 | 0.681 |

OR: Odds Ratio; CI: Confidence Interval; RM: Regional median mortality rate=118.45 per 1000 live births

### Universal Health Coverage for Under-five Children

More than two thirds of urban HDs recorded a proportion of free malaria treatment cases below the regional median (14%), with the lowest rates found in Nkolndongo and Odza HDs. In rural areas, one third of HDs performed below this regional threshold. The worst performing health districts were Mbalmayo, Ndikinimeki, Mbandjock and Okola (Figure 9, Table 6).

**Fig. 9.**
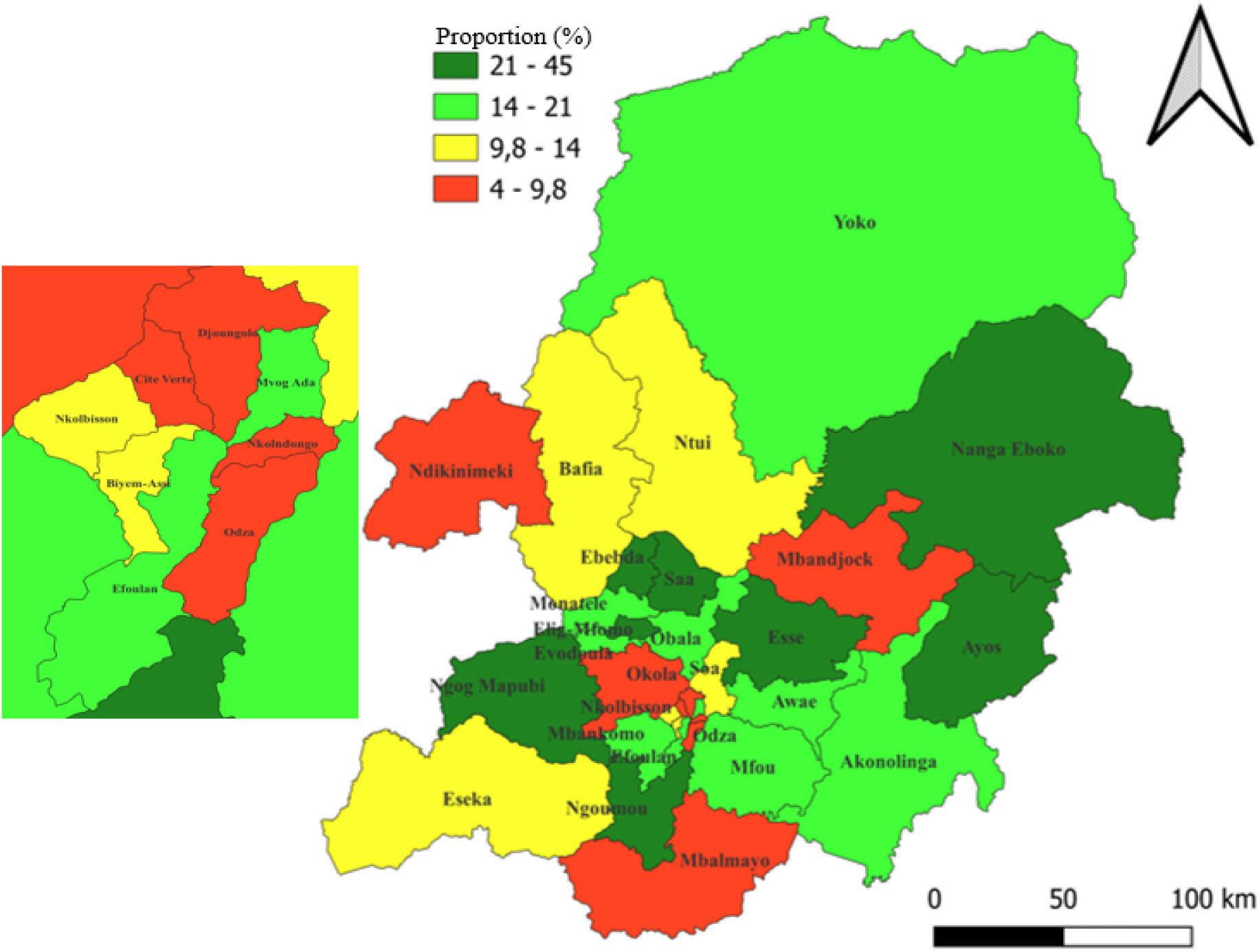
Proportion of malaria cases treated free of charge in Centre Region Health Districts from 2018 to 2022

**Table 6.** Association between the type of environment and poor compliance with the free malaria treatment policy for under five children in the Centre Region (*n*=32)

| Environment | Mortality rate<br>*n* (%) |  | Total<br>(100%) | Association test |  |  |
| --- | --- | --- | --- | --- | --- | --- |
| | ≥ RM<br>*n*=18 | < RM<br>*n*=14 | | OR | 95% CI | *p*-value |
| Rural | 16 (66.7) | 8 (33.3) | 24 | 1 |  |  |
| Urbain | 2 (25.0) | 6 (75.0) | 8 | 1.41 | 0.28-8.16 | 0.681 |

OR: Odds Ratio; CI: Confidence Interval; RM: Regional median proportion=14%

## Discussion

Human resources for health in the Centre Region remain well below the WHO standard of 23-25 qualified health workers per 10,000 inhabitants to respond more effectively to people’s health problems [13,15]. This observation was highlighted in a national assessment of Cameroon in 2013 [16]. Key challenges in the country’s human resource management system include the absence of a recruitment plan, low investment from the private sector, and a disconnect between health worker training and employment opportunities [17].

The Region’s inpatient mortality rates were evenly distributed across the areas. The approach to reducing hospital deaths is based on aspects such as the density of qualified human resources for health and the pricing of care. In a context of lack of health insurance, the cost of care is often a barrier to patients’ access to quality care in both public and private HFs, especially in emergency situations. In addition, improving the safety of hospital care and reducing hospital deaths is a clear goal that is well supported by clinicians, managers and patients. Good leadership and information, a quality improvement strategy based on good local evidence and a community-wide approach can be effective in improving the quality of care processes and reducing hospital mortality [18].

A higher risk of perioperative mortality was observed in rural areas compared with urban areas. The quality of care in the management of emergencies, especially surgical emergencies, hospital hygiene and the level of postoperative follow-up may explain these findings. Urban areas have a greater number of specialists and a better standard of care than rural areas [16]. The humanization of hospital care includes high quality care and better management of patients and their families, especially in emergency situations. It is therefore necessary to carry out surveys in rural district HFs to ensure that these care meet hygiene standards. However, the lack of use of a surgical checklist, patient comorbidities, blood transfusions and the use of general anesthesia are predictors of death in healthcare settings [19].

Neonatal mortality was higher than the regional average in most urban HDs, and maternal mortality remained a major problem in remote rural HDs. This situation can be explained, on the one hand, by the inadequate implementation of public health policies on maternal and neonatal health in some of the region’s health districts and, on the other, by the poor compliance of the population with these general measures. In fact, it is possible to improve neonatal survival and health and eliminate avoidable stillbirths by achieving high coverage of quality antenatal care, skilled care at birth, postnatal care for mother and child, and care for small and sick newborns [20].

The death cases observed in the Region may be attributed to various factors. The rate of use of hospital care was 10 times lower than the recommended standard (Table 1). This low level of use results in delayed treatment, thereby increasing the risk of mortality. In addition, self-medication, exacerbated by the illicit sale of street drugs, financial constraints linked to the unfavorable economic context and the absence of a universal health coverage mechanism at that period, can have varying degrees of impact on patient survival [21,22].

Most urban HDs did not comply with the national malaria treatment policy for children under five. The high demand for drugs compared to supply in urban HFs may explain this poor performance. Indeed, these HFs are constantly experiencing stock-outs of inputs such as artemether-lumefantrine and injectable artesunate, which are needed to implement this health coverage. However, the introduction of universal health coverage in the country could help to address this health issue, by ensuring a better monitoring of hospital supplies of free malaria treatment inputs [23–25].

## Limitations

We used secondary data for which the quality of the collection and transmission process could not be fully guaranteed.

## Conclusions

Humanization of health care in the Centre Region remains a challenge for the health authorities, as inequalities persist in terms of the quality of care and the effective implementation of health policies between urban and rural HDs. This study has enabled us to identify the priority HDs with alarming indicators in terms of the quality of hospital care, in a context of a shortage of qualified health professionals in both urban and rural areas. In urban areas, alerts were identified in the quality and safety of hospital care in general and in the implementation of the national policy for the prevention of neonatal mortality in particular. On the other hand, in rural HDs, warning signs were identified in surgical care and hospital care related to maternal health. Better compliance with the free malaria treatment policy could reduce the financial burden of this disease on families, especially in urban areas, and ensure that children under five are safe from malaria. Effective implementation of universal health coverage will help pave the path for achieving the third objective of SDGs.

## Recommendations

To address the various challenges identified in this study, we suggest that the health authorities to mobilize fund for the implementation of the National Strategic Plan to Improve the Quality of Health Care and Services through training and supervision; propose strategies to redeploy and ensure the retention of doctors and specialists in priority areas; organize further operational research to identify the root causes of the problems identified; and ensure a continuous supply of free malaria treatment commodities.

## Data Availability

All data produced in the present work are contained in the manuscript

## Abbreviations

DHIS: District Health Information System
HCW: Healthcare Worker
HD: Health District
HF: Health Facility
SDG: Sustainable Development Goal

## Declarations

### Authors’ Contribution

Study design & conception: FZLC; Data collection and processing: FZLC and CM; Data analysis, visualization and interpretation: FZLC; Drafting of original manuscript: FZLC; Critical revision of the manuscript: FZLC, AA, BNA, LBKB, CM, MGMT, EABBM, AN, CSN, CAM, EOG, RKNO, D-H, AY, PS and FKN; Final approval of the manuscript: All authors.

### Ethical Approval Statement

Ethical clearance for the present study was waived by the Faculty of Medicine of Yaounde’s ethical review board. As this secondary data was anonymous and publicly available in aggregated form on the DHIS2 platform, there was no risk to the individuals whose data was collected. All methods were performed in accordance with the relevant guidelines of the Helsinki declaration.

### Consent for publication

Not applicable.

### Availability of data and materials

All data generated or analyzed during this study are included in this published article.

